# A 10-Year Clinico-Epidemiological Study of Snake Bite Cases in Siraha District

**DOI:** 10.1101/2025.10.13.25337869

**Authors:** Akash Raut, Puja Sah, Suraj Shrestha, Sabin Thapaliya

## Abstract

**Background:** Snakebite envenoming is a major public health concern in Nepal, particularly in agricultural regions such as Siraha District. It disproportionately affects rural populations, where delayed hospital presentation and harmful first-aid practices contribute to preventable morbidity and mortality. Despite global WHO strategies to reduce the snakebite burden by 2030, comprehensive long-term data from Nepal remain limited.

**Methods:** A retrospective descriptive study was conducted at Ram Kumar Sarda Uma Prasad Murarka Provincial Hospital, Lahan, covering ten years (2071–2080 BS / 2014–2024 AD). All clinically confirmed or strongly suspected snakebite cases were included. Data on demographics, bite characteristics, clinical features, prehospital interventions, treatments, and outcomes were extracted from hospital records and analyzed using SPSS version 25. Annual case fatality rates (CFR) were calculated.

**Results:** A total of 3,499 snakebite cases were recorded, with 58 deaths (CFR 1.66%). Adults aged 16–45 years (60.5%) and farmers were most affected. Most bites occurred on lower limbs (67.2%), during evening hours and the monsoon season (44.3%). Only 3.4% of patients presented within one hour, while 38.2% arrived after six hours. Harmful first-aid practices particularly tourniquet use (17.6%) and local incisions (3.9%) remained common. Cytotoxic manifestations predominated, while severe systemic symptoms were infrequent.

**Conclusion:** Snakebite remains a significant occupational and seasonal hazard in Siraha District. Although the low CFR indicates improved case management, delayed presentation and unsafe prehospital care persist. Strengthening community education, ensuring early access to antivenom, and improving diagnostic and referral systems are essential to further reduce mortality and disability.

## Introduction

Snakebite envenoming represents a significant global public health challenge, causing between 81,000 and 138,000 deaths annually worldwide, with an additional 400,000 amputations and other permanent disabilities [1]. This neglected tropical disease disproportionately affects rural populations in low- and middle-income countries, particularly agricultural communities in South Asia, sub-Saharan Africa, and parts of Latin America [2]. Recognizing this burden, the World Health Organization (WHO) included snakebite envenoming in its list of neglected tropical diseases in 2017 and subsequently released a comprehensive global strategy in 2019 to halve snakebite mortality and morbidity by 2030 [3].

South Asia bears the heaviest burden of snakebite envenoming globally, contributing a majority of snakebite-related deaths [4]. Within this region, Nepal faces a particularly challenging situation, with WHO estimates indicating approximately 20,000 snakebite cases annually resulting in over 1,000 deaths [5]. However, these figures likely underestimate the true burden, as hospital- and community-based studies from Nepal have suggested substantially higher incidence rates [6,7].

The epidemiological profile of snakebite in Nepal mirrors broader regional patterns, with peak incidence during monsoon months when increased agricultural activity and flooding drive snakes into human settlements [7,8]. Hospital-based studies across different regions have consistently demonstrated significant case fatality rates, influenced by healthcare infrastructure, antivenom availability, and treatment protocols [9–13]. The demographic profile typically shows predominance among young adults aged 16–45 years, particularly farmers and agricultural workers, reflecting occupational exposure risks [9,12,13].

Critical gaps in emergency care delivery remain a persistent challenge throughout Nepal’s healthcare system. Delayed hospital presentation is common, with significant proportions of patients arriving more than six hours post-bite, beyond the optimal window for antivenom effectiveness [14–18]. Traditional first aid practices, including tourniquet application and local incision, are still widespread and may exacerbate tissue damage and systemic complications [12,19,20]. Timely antivenom administration is crucial, as delays beyond six hours significantly increase mortality and treatment complexity [15,16,18].

Clinically, snakebite envenoming in Nepal predominantly reflects cytotoxic manifestations consistent with regional viper species, though neurotoxic presentations from elapid species also occur [13,16,21]. Species identification remains challenging, as most cases involve unidentified snakes, limiting opportunities for species-specific management [17]. This is further compounded by limited laboratory capacity and inadequate training in clinical assessment of envenomation severity [13].

Recent advances in snakebite management, including updated WHO guidelines and improved antivenom formulations, offer promise for reducing mortality and morbidity [16,19]. Successful implementation requires robust surveillance systems, adequate healthcare infrastructure, and community education programs to address knowledge gaps and harmful practices [24,25]. The establishment of dedicated snakebite treatment centers and healthcare worker training programs has demonstrated encouraging results in reducing case fatality rates in some regions [19,23].

This study aims to provide comprehensive epidemiological insights into snakebite patterns at Provincial Hospital Lahan, Siraha district, over a ten-year period from 2071–2080 BS (2014–2024 AD). By analyzing demographic characteristics, seasonal patterns, clinical presentations, treatment outcomes, and healthcare delivery factors, this research seeks to inform evidence-based strategies for snakebite prevention and management in Nepal’s Terai region, contributing to the achievement of WHO’s 2030 goals for snakebite burden reduction [3].

## Methodology

### Study Design

This was a retrospective descriptive hospital-based study conducted over a 10-year period, from Baisakh 2071 to Chaitra 2080 B.S. (April 2014 to March 2024 A.D.). The study aimed to describe the clinico-epidemiological profile, temporal patterns, and treatment outcomes of snakebite cases managed at a tertiary-level hospital in southeastern Nepal.

### Study Setting

The study was carried out at Ram Kumar Sarda Uma Prasad Murarka Provincial Hospital, Lahan, situated in Siraha District, Madhesh Province, Nepal.

The hospital serves as a major referral center for Siraha and adjoining districts and provides both emergency and inpatient services. The district’s predominantly agricultural and rural population faces a high risk of snakebite envenomation, particularly during the monsoon and post-harvest seasons when human-snake encounters are frequent.

### Study Population

The study included all patients presenting with a history or clinical diagnosis of snakebite during the specified 10-year period.

Cases were identified through emergency and inpatient records, antivenom (ASV) logs, and hospital case files.

### Inclusion Criteria

- All patients of any age and gender presenting with a clinically confirmed or strongly suspected snakebite.
- Records containing complete demographic, clinical, treatment, and outcome details.
- Cases documented in official hospital registers, including emergency, inpatient, or ASV distribution logs.

### Exclusion Criteria

- Records with incomplete or missing key variables (age, gender, clinical findings, or outcomes).
- Non-snakebite envenomation cases (e.g., scorpion or animal bites).
- Duplicate entries or cases lacking adequate confirmation of snakebite exposure.

### Data Collection Procedure

Data were collected retrospectively from hospital archives using a pre-designed structured data extraction form.

Variables included:

- **Demographic details:** age, gender, and occupation.
- **Bite characteristics:** anatomical site, time of bite, season, and snake species (if identified).
- **Clinical features:** local and systemic manifestations of envenomation.
- **Prehospital interventions:** first aid measures, including traditional or harmful practices (tourniquet, incision, herbal remedies).
- **Treatment-related variables:** time interval between bite and hospital arrival, administration of antivenom (ASV), requirement of intensive care unit (ICU) admission, dialysis, or blood transfusion.
- **Outcome variables:** survival, complications, or death.

### Data Management and Analysis

Data were compiled in Microsoft Excel 2021 and analyzed using IBM SPSS Statistics version25. Descriptive statistics were computed, including frequencies, percentages, means, and standard deviations for quantitative variables. Year-wise and seasonal temporal trends were represented through line and bar graphs. Comparative analyses were performed to explore associations between demographic variables, time of presentation, first aid practices, and clinical outcomes. The Case Fatality Rate (CFR) was calculated annually as the proportion of snakebite-related deaths to total cases in that year.

Ethical approval was obtained from the Institutional Review Committee of National Health Research Council, Nepal, and all patient identifiers were removed to maintain confidentiality.

## Results

### Study Population and Temporal Trends

Between April 2014 and March 2024 (Baisakh 2071–Chaitra 2080), Provincial Hospital Lahan recorded 3,499 snakebite cases, with 58 deaths (case fatality rate [CFR] 1.66%). Annual cases ranged from 286 to 437, peaking in 2071 and lowest in 2076. Mortality rates varied between 0.90% and 2.46% over the study period. The mean annual number of cases was 349.9 (SD 43.0), with an average of 5.8 deaths per year (SD 1.8).

**Fig. 1.**
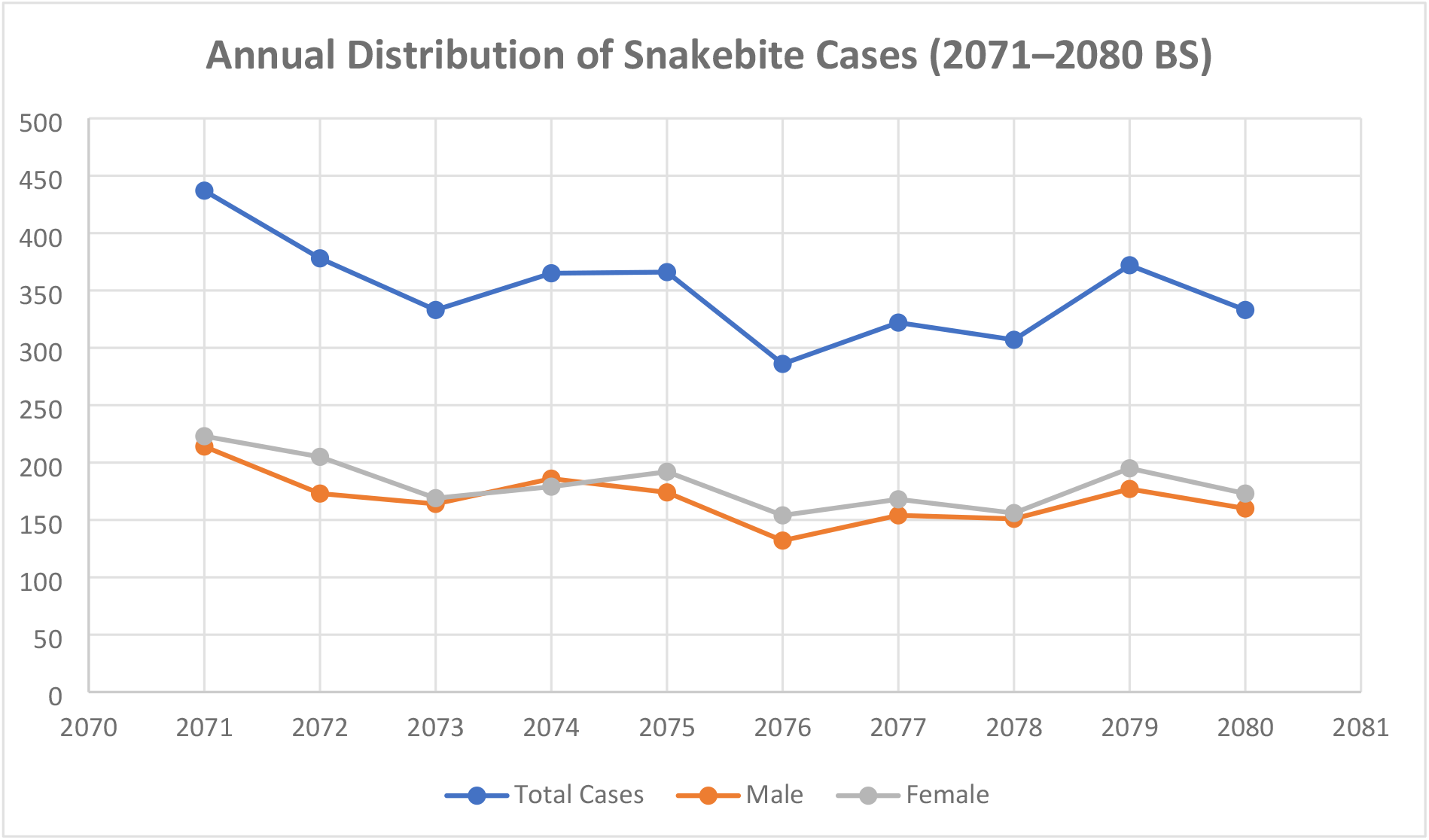
Annual Distribution of Snakebite Cases (2071–2080 BS)

### Demographic Characteristics

Adults aged 16–45 years bore the highest burden (60.5%), with 16–30 years comprising 33% and 31–45 years 27.5%. Children (0–15 years) accounted for 19.3%, those aged 46–60 years 18.3%, and >60 years 2.3%. Females consistently outnumbered males across all years.

### Anatomical distribution of snake bite site

**Fig. 2.**
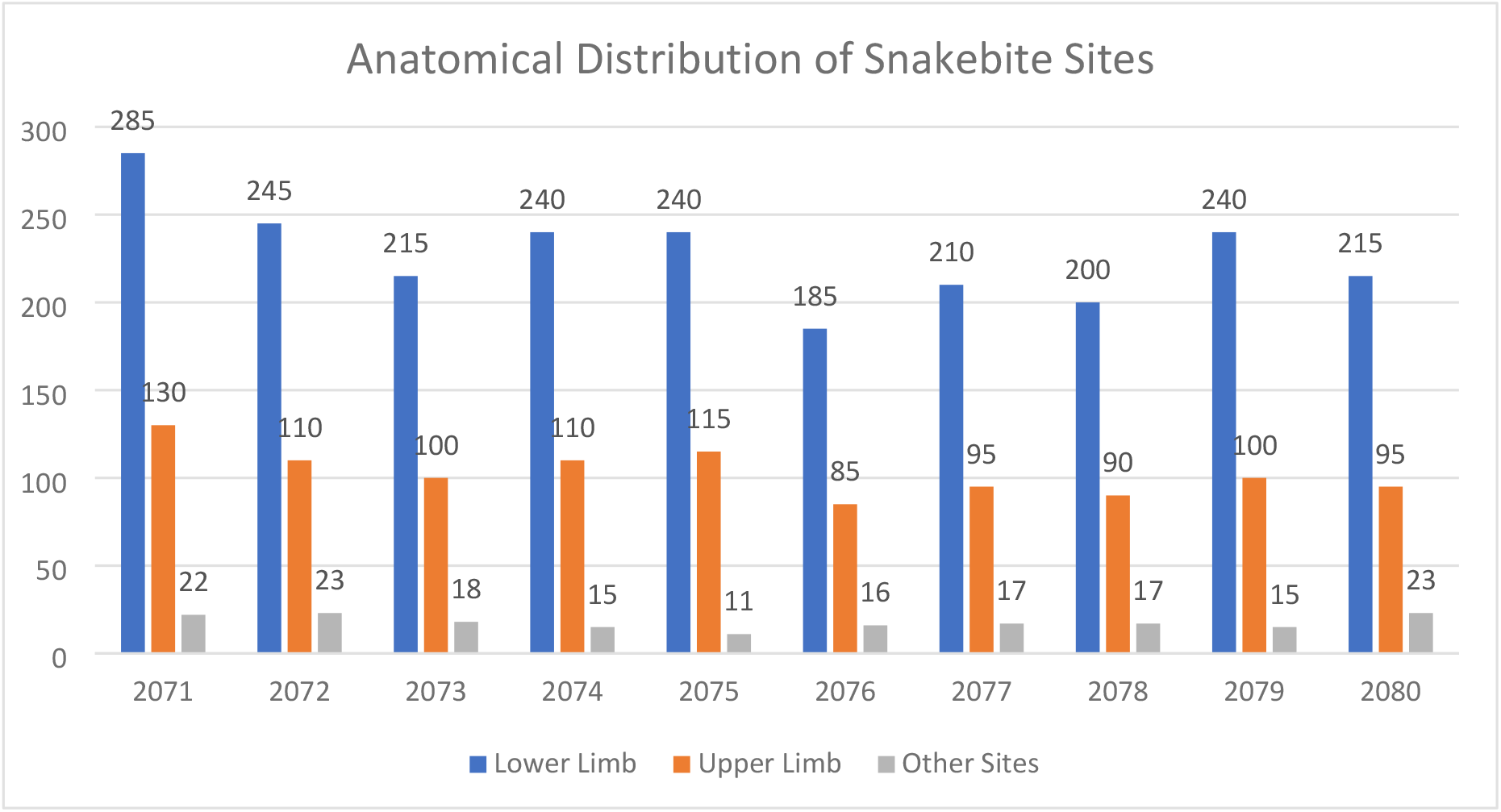
Anatomical Distribution of Snakebite Sites

The predominance of lower limb involvement (67.2%) is expected in rural Nepal, where most bites occur during farming, walking barefoot, or during evening hours.Upper limb involvement (30.4%) likely results from manual work, handling firewood, or attempting to catch or remove the snake. The low proportion of “other sites” (5.2%) may indicate that bites during sleep or surprise encounters were less common, but still notable.The year 2076 BS shows the lowest site-specific cases across all locations, which aligns with the lowest total snakebite cases that year (286 cases).

### Hospital Presentation and Treatment-Seeking Behavior

Only 3.4% of patients presented to the hospital within one hour of the bite. The majority arrived within 1–6 hours (58.4%), while 38.2% sought care after more than six hours, including 12.2% after 12 hours.

### First Aid Practices

Most victims (58%) received no first aid. Harmful practices were common: tourniquet application (17.6%), traditional/herbal remedies (7.8%), and local incisions (3.9%). Proper immobilization, consistent with recommended guidelines, was documented in 5.5% of cases, with gradual improvement over the decade.

### Clinical Presentations

Pain was the most frequent symptom (90.2%). Local complications included cellulitis (42.7%), bleeding (28%), and gangrene (4.3%). Systemic manifestations were less common: nausea (7.5%), vomiting (6.4%), ptosis (4.1%), dysphagia (1.8%), respiratory distress (1.6%), unconsciousness (1.0%), and convulsions (0.7%).

**Table 1.** Distribution of Clinical Symptoms in Snakebite Cases (2071–2080 BS)

| Year | Total Cases | Pain | Bleeding | Cellulitis | Gangrene | Nausea | Vomiting | Ptosis | Resp. Distress | Dysphagia | Unconsciousness | Convulsions |
| --- | --- | --- | --- | --- | --- | --- | --- | --- | --- | --- | --- | --- |
| 2071 | 437 | 390 | 120 | 200 | 20 | 30 | 25 | 15 | 5 | 7 | 3 | 2 |
| 2072 | 378 | 340 | 110 | 180 | 18 | 28 | 22 | 14 | 6 | 6 | 4 | 3 |
| 2073 | 333 | 310 | 90 | 150 | 15 | 25 | 20 | 13 | 4 | 5 | 3 | 2 |
| 2074 | 365 | 320 | 100 | 170 | 17 | 29 | 24 | 16 | 6 | 7 | 5 | 3 |
| 2075 | 366 | 325 | 105 | 175 | 19 | 35 | 30 | 18 | 8 | 9 | 6 | 4 |
| 2076 | 286 | 260 | 85 | 130 | 12 | 20 | 18 | 12 | 4 | 4 | 2 | 2 |
| 2077 | 322 | 295 | 90 | 140 | 14 | 22 | 20 | 13 | 5 | 5 | 3 | 2 |
| 2078 | 307 | 280 | 85 | 135 | 10 | 21 | 19 | 12 | 4 | 4 | 2 | 1 |
| 2079 | 372 | 335 | 100 | 165 | 15 | 27 | 25 | 16 | 6 | 6 | 4 | 2 |
| 2080 | 333 | 300 | 95 | 150 | 11 | 24 | 22 | 14 | 4 | 5 | 2 | 2 |

**Fig. 3.**
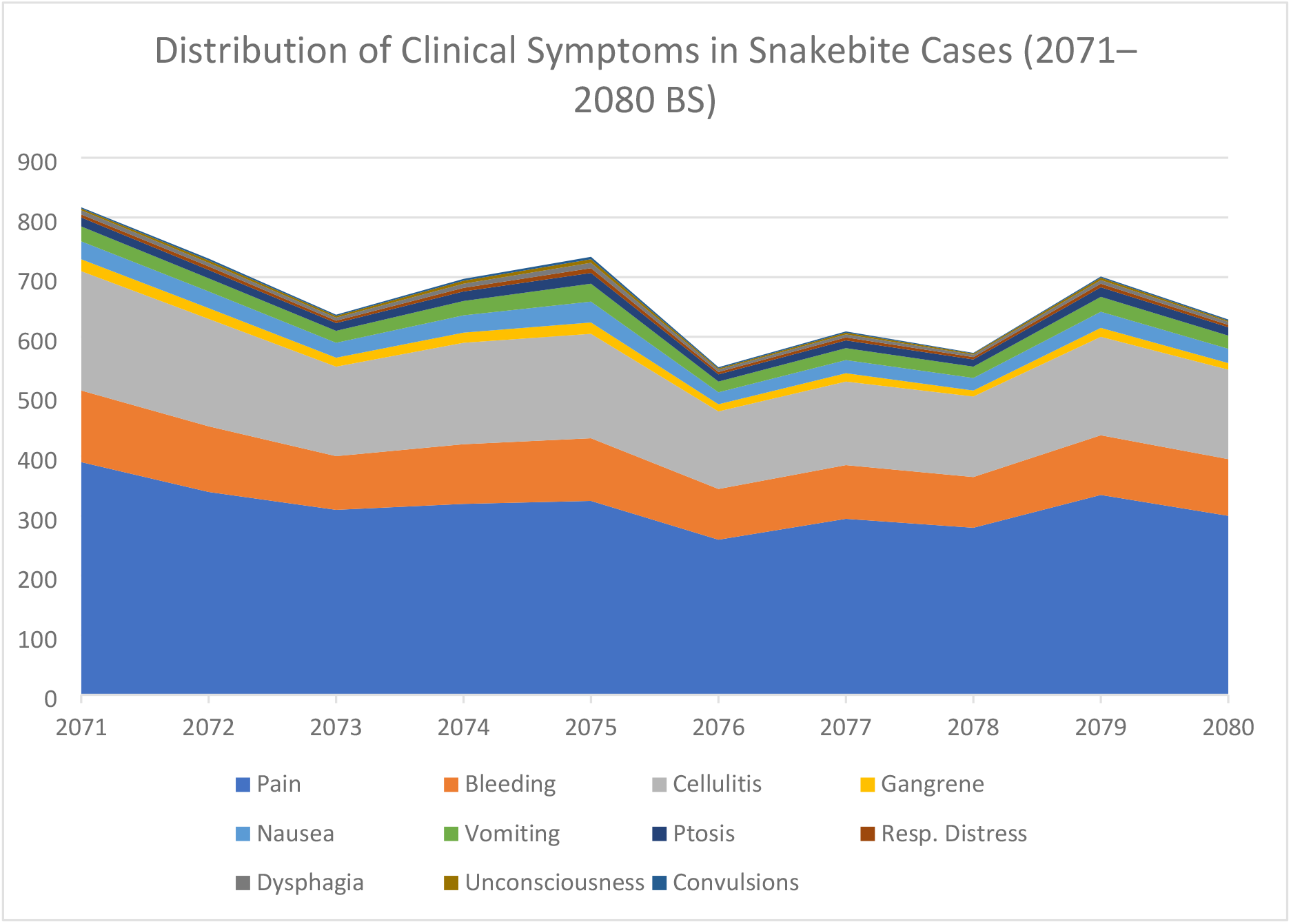
Distribution of Clinical Symptoms in Snakebite Cases (2071–2080 BS)

### Occupational Distribution

Farmers were the most affected group, followed by students and housewives. Laborers and other occupations accounted for fewer cases. Peak occupational risk was observed in 2071 (Farmers: 200; Students: 110) and the lowest in 2076 (Farmers: 140; Students: 70).

### Snake Species

Most bites were from unidentified species. Documented cobra bites ranged from 3 to 7 annually, while krait bites were rare (0–3 per year). The number of bites by unknown species declined from 432 in 2071 to 276 in 2076, with minor fluctuations thereafter.

### Time of Bite

Snakebites occurred most frequently in the evening (4–8 PM), followed by the afternoon (12–4 PM). Morning (6 AM–12 PM) and night (8 PM–6 AM) periods recorded fewer cases.

### Seasonal Distribution

Incidence peaked during the monsoon (Shrawan–Bhadra, 44.3%), followed by summer (Jestha– Ashad, 30.2%). Autumn (Ashoj–Kartik) and spring (Chaitra–Baisakh) contributed 12.7% and 9.4%, respectively, while winter (Mangsir–Falgun) had the lowest incidence (3.4%).

**Table 2.** Seasonal and Monthly Trend of Snake Bite Incidence (2071–2080 B.S.)

| Year | Spring<br>(Chaitra–<br>Baisakh) | Summer<br>(Jestha–<br>Ashad) | Monsoon<br>(Shrawan–<br>Bhadra) | Autumn (Ashoj–<br>Kartik) | Winter<br>(Mangsir–<br>Falgun) |
| --- | --- | --- | --- | --- | --- |
| 2071 | 40 | 125 | 180 | 55 | 37 |
| 2072 | 33 | 110 | 160 | 50 | 25 |
| 2073 | 28 | 95 | 145 | 40 | 25 |
| 2074 | 30 | 105 | 160 | 45 | 25 |
| 2075 | 35 | 110 | 165 | 40 | 16 |
| 2076 | 25 | 90 | 130 | 35 | 6 |
| 2077 | 30 | 95 | 140 | 38 | 19 |
| 2078 | 29 | 100 | 135 | 34 | 9 |
| 2079 | 38 | 115 | 150 | 42 | 27 |
| 2080 | 35 | 105 | 145 | 38 | 10 |

**Fig. 4.**
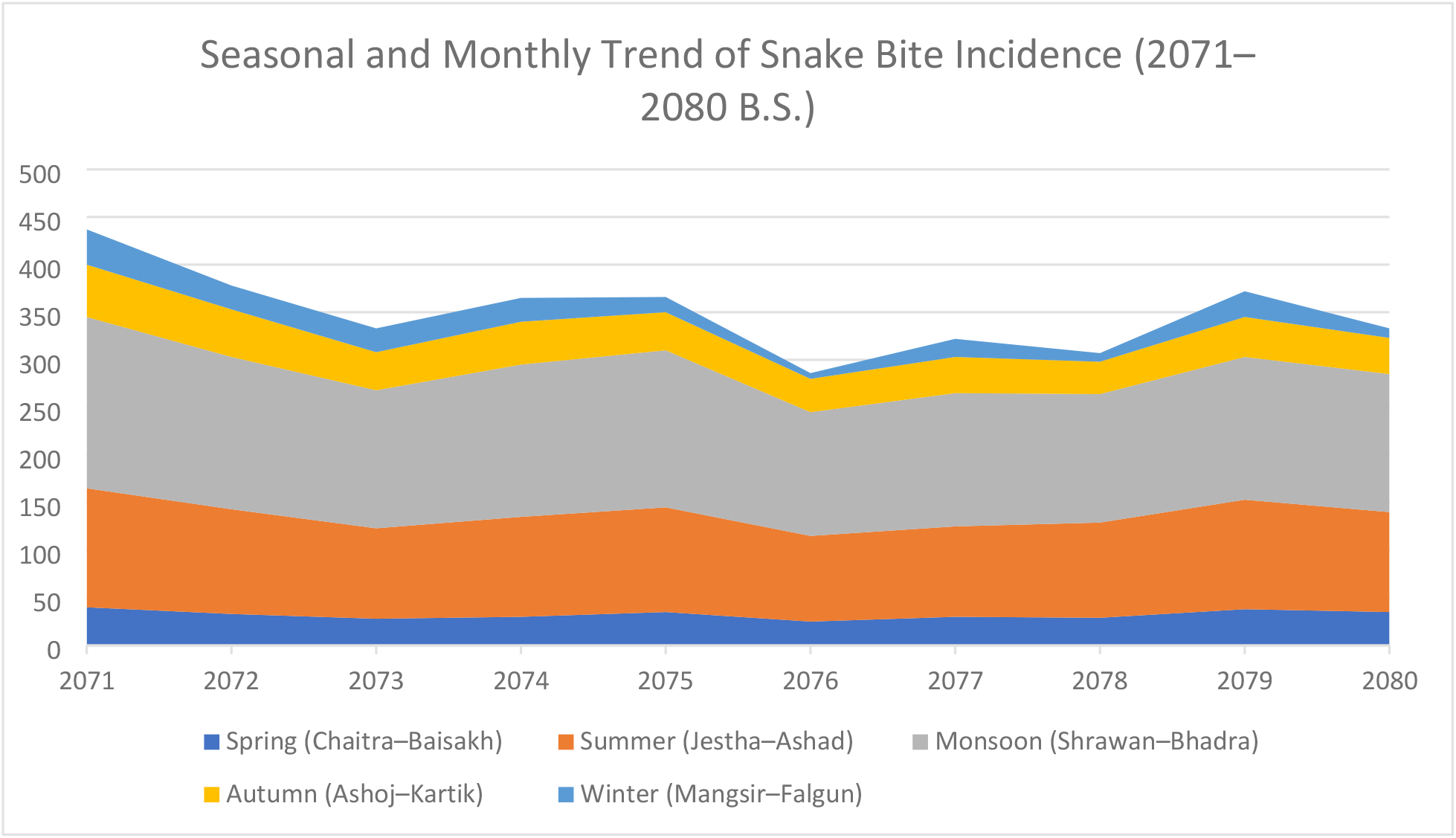
Seasonal and Monthly Trend of Snake Bite Incidence (2071–2080 B.S.)

### Treatment Outcomes

The overall CFR was 1.66%, lower than previously reported in Nepal (3–58%). Annual mortality fluctuated between 0.9% and 2.46%, peaking in 2075 and reaching the lowest in 2080. Delayed hospital presentation (>6 hours) remained a key determinant of adverse outcomes.

### Therapeutic Interventions for Snakebite Envenomation

ASV usage does not directly correlate with total case load, but slightly increases in years with more complicated cases.2075 BS stands out with the highest use of ASV, diuretics, blood transfusions, dialysis, and ICU admissions, despite only moderate total case numbers— suggesting greater case severity or better reporting/intervention access.A generally low rate of advanced intervention (e.g., dialysis, ICU) reflects that most snakebites were likely non-severe or dry bites, or that referral/ICU capacity was limited.

**Table 3.** Therapeutic Interventions for Snakebite Envenomation.

| Year | Total Cases | ASV Given | Diuretics | Hemodialysis | Blood Transfusion | ICU Admissions |
| --- | --- | --- | --- | --- | --- | --- |
| 2071 | 437 | 30 | 10 | 2 | 5 | 3 |
| 2072 | 378 | 28 | 9 | 3 | 4 | 4 |
| 2073 | 333 | 26 | 8 | 2 | 4 | 3 |
| 2074 | 365 | 32 | 11 | 4 | 6 | 5 |
| 2075 | 366 | 35 | 13 | 5 | 7 | 6 |
| 2076 | 286 | 22 | 7 | 2 | 3 | 2 |
| 2077 | 322 | 24 | 9 | 3 | 4 | 3 |
| 2078 | 307 | 26 | 10 | 3 | 5 | 4 |
| 2079 | 372 | 29 | 11 | 4 | 6 | 5 |
| 2080 | 333 | 25 | 9 | 3 | 4 | 3 |

**Fig. 5.**
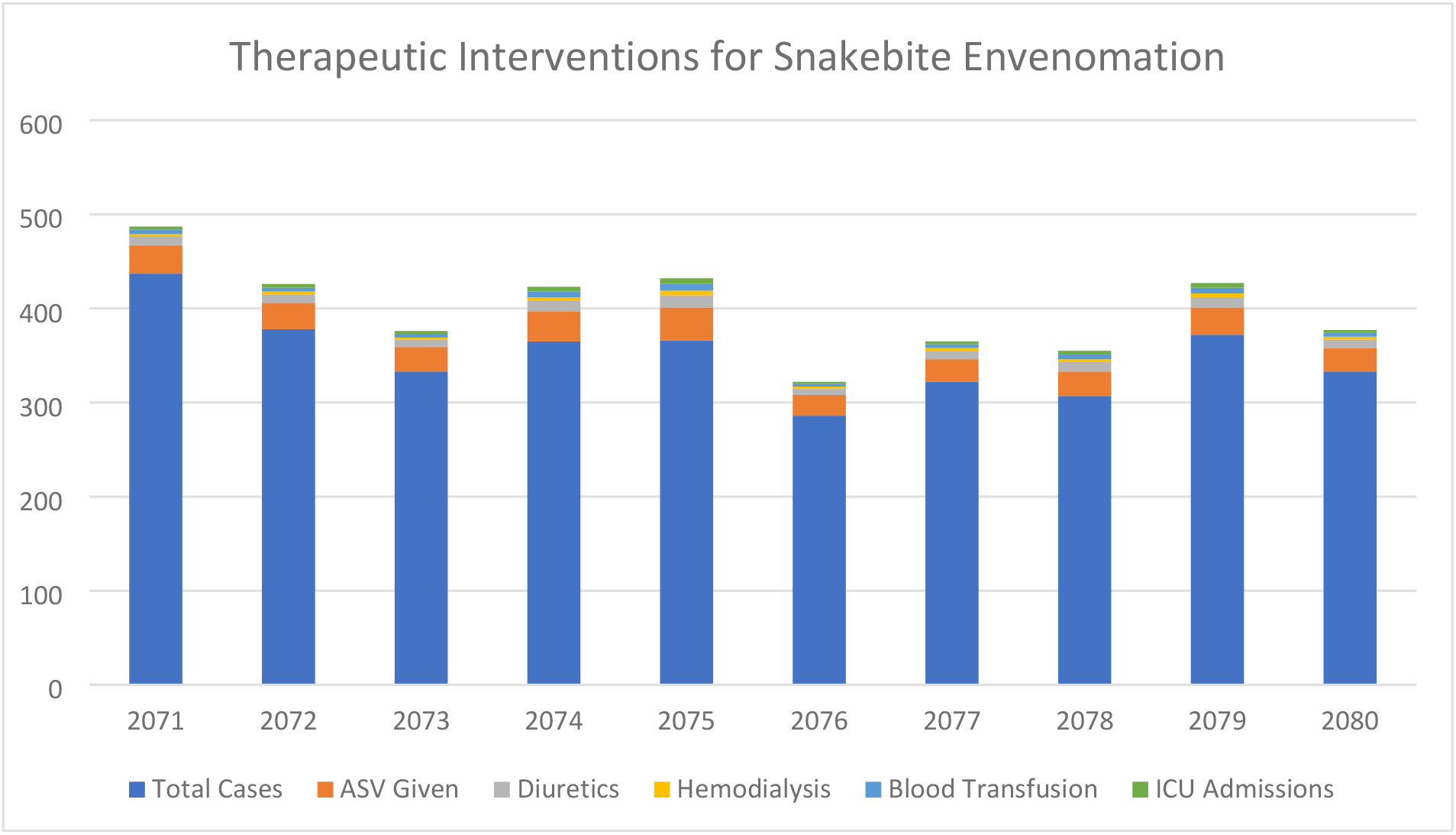
Therapeutic Interventions for Snakebite Envenomation

## Discussion

This ten-year clinico-epidemiological analysis of 3,499 snakebite cases at Provincial Hospital Lahan reveals several important findings aligning with broader South Asian patterns while demonstrating unique regional characteristics. The case fatality rate of 1.66% represents a substantial improvement compared to earlier Nepalese studies reporting mortalities ranging from 3–58% [1,6,7,11], suggesting enhanced case management protocols, improved antivenom availability, and better healthcare infrastructure at the district level [5,14,18,19,20].

The demographic profile confirms snakebites as primarily an occupational hazard, with adults aged 16–45 years constituting 60.5% of cases, particularly affecting farmers who represented the largest occupational group throughout the study period. This finding corresponds with established patterns across South Asia, where agricultural communities face the highest burden [1,4,7,15,20]. The notable pediatric burden of 19.3% aligns closely with previous studies reporting children accounting for 25% of cases in Nepal [2,3,13], with higher vulnerability attributed to behavioral factors including outdoor exploration, barefoot walking, and increased exposure during agricultural activities [13].

Seasonal patterns strongly reflect monsoon-driven epidemiology, with 44.3% of cases occurring during Shrawan–Bhadra months, consistent with established South Asian trends where flooding forces snakes into human settlements [4,7,15]. The temporal distribution showing peak incidence during evening hours (4–8 PM) mirrors findings from other Nepalese studies, reflecting periods of maximum outdoor activity and reduced visibility [6,7,11].

Critical gaps in emergency care remain evident, with only 3.4% of patients reaching medical facilities within one hour and 38.2% presenting beyond six hours post-bite. This delay pattern significantly impacts antivenom effectiveness, as multiple studies demonstrate optimal outcomes when administered within six hours [8,12,19]. The predominance of harmful first aid practices, including tourniquets (17.6%) and local incisions (3.9%), reflects persistent community misconceptions that may worsen outcomes [9,17,20], emphasizing the need for targeted education campaigns.

The clinical presentation pattern, dominated by cytotoxic manifestations (cellulitis 42.7%, bleeding 28.0%) rather than neurotoxic symptoms, suggests regional predominance of viperid species consistent with Terai ecology [4,6,7,11]. The high proportion of unidentified species (89– 95% across years) highlights ongoing diagnostic challenges that impact species-specific management decisions [5,14,18,20].

The study’s limitations include single-center design potentially limiting generalizability, lack of species confirmation affecting targeted treatment approaches, and hospital-based data that may underestimate community burden [11,15,20]. However, the large sample size and decade-long duration provide valuable insights into temporal trends and clinical outcomes.

The favorable mortality outcome compared to historical data suggests potential effectiveness of current treatment protocols [1,6,11,18,20], yet the persistent burden of delayed presentation and harmful first aid practices indicates substantial room for improvement [9,17,19]. Future interventions should focus on community education programs to promote appropriate first aid measures, establishment of peripheral antivenom stockpiles to reduce treatment delays [5,14,19,20], and implementation of rapid diagnostic tools for species identification to optimize clinical management [5,14,18,20].

## Conclusion

This ten-year retrospective study highlights snakebite envenoming as a persistent public health challenge in southeastern Nepal, predominantly affecting young adults and farmers during the monsoon season. Although the case fatality rate of 1.66% indicates improved hospital management, delayed presentation and harmful first-aid practices remain major concerns. The predominance of cytotoxic features suggests regional viperid dominance, while the high proportion of unidentified species emphasizes the need for improved diagnostic capacity. Expanding community education, ensuring early access to antivenom at peripheral centers, and strengthening surveillance and clinical training are critical to further reduce mortality and morbidity.In conclusion, sustained multisectoral efforts integrating community awareness, rapid referral, and evidence-based treatment can substantially reduce the burden of snakebite in Nepal and contribute toward achieving the WHO 2030 goal of halving global snakebite deaths and disabilities.

## Data Availability

All relevant data underlying the findings of this study are included within the manuscript and its Supporting Information files. Additional de-identified data from hospital records can be made available upon reasonable request to the corresponding author, subject to approval by the Institutional Review Board of Ram Kumar Sarda Uma Prasad Murarka Provincial Hospital, Lahan, to ensure patient confidentiality

